# Psychiatric Residential Treatment Facilities for Child Behavioral Health Services in North Carolina Medicaid

**DOI:** 10.1101/2022.12.21.22283789

**Authors:** Paul Lanier, Roderick Rose, Daniel Gibbs, Jacob Hyman, Neil Kamdar, Joseph Konstanzer, Kristen Hassmiller Lich

## Abstract

**Background:** Psychiatric residential treatment facilities (PRTFs) are a type of non-hospital inpatient treatment setting for children with severe behavioral health disorders. PRTFs are a restrictive and costly form of care that can potentially be avoided with community-based behavioral health services.

**Methods:** Statewide Medicaid enrollment and claims data for 2015 to 2022 were used to describe PRTF utilization in North Carolina. We examined annual episodes of care in PRTFs and compared trends before and during the COVID-19 public health emergency.

**Results:** From 2015 to 2022, 10,038 children insured by North Carolina Medicaid entered a PRTF across 10,966 episodes of care. In the past five years (2018-2022), care in PRTFs resulted in Medicaid expenditures of over $550 million total, or over $100 million per year. In 2022, 42% of children who entered PRTFs were in foster care and 44% of children were placed in PRTFs outside of North Carolina.

**Limitations:** Analysis limited to data collected for administrative purposes.

**Conclusions:** Current trends indicate ongoing overrepresentation of children in foster care placed in PRTFs and increased out-of-state PRTF placements. Coordinated efforts in future research, policy, and practice are needed to determine the cause of these trends and identify solutions.

## Introduction

Over 1 in 5 children in North Carolina (NC) – about 450,000 children statewide – have a mental, emotional, developmental, or behavioral health problem.^1^ Federal policy requires that children receive behavioral health services and education in the least restrictive setting possible within their own community.^2^ Although many behavioral health challenges can be adequately treated with community-based outpatient services, some children with severe symptoms may require supervised restrictive care.

Psychiatric residential treatment facilities (PRTFs) are defined by the Centers for Medicaid and Medicare Services as a “non-hospital facility with a provider agreement with a State Medicaid Agency to provide the inpatient services benefit to Medicaid-eligible individuals under the age of 21.”^3^ In NC, children referred to PRTFs must meet the criteria outlined in the NC Division of Health Benefits’ certificate of need^4^: (a) the community’s ambulatory care resources do not meet the treatment needs of the recipient; (b) proper treatment of the recipient’s condition requires services on an inpatient basis overseen by a physician; (c) inpatient services can reasonably be expected to improve the recipient’s condition or prevent further regression so that services will no longer be needed. These criteria ensure that children are only placed in PRTFs because their needs cannot be met in another treatment setting.

Children with mental health challenges are also more likely to come in contact with the child welfare system and to enter foster care.^5,6^ Because maltreated children are more likely to need mental health services^7,8^, it is understandable that children involved with child welfare are overrepresented in PRTFs. However, when less restrictive child welfare placements (e.g., foster care, kinship care) are unavailable, restrictive settings such as PRTFs are often used as a stable placement, regardless of whether the child’s care medically requires this setting. Because Black and Native American children are overrepresented in the child welfare system^9^, understanding the connection between the child welfare system and PRTFs is crucial to promoting racial equity – a stated commitment of the NC Department of Health and Human Services.^10^

Although federal and state policies state that PRTFs should only be used when medically necessary, advocacy groups have raised concerns about the financial and ethical costs of overutilizing PRTFs.^11^ Concerningly, children already in foster care placement are at highest risk of unnecessary placement in restrictive care. Thus, while overuse of PRTFs indicates a clear clinical need in a population, it also reflects that population’s poor access to quality community-based services. Monitoring PRTF utilization rates and trends is crucial to ensuring appropriate treatment is available to children, particularly those already in contact with the child welfare system.

### National Studies

PRTFs’ national usage rates are neither regularly monitored nor comprehensively understood. A 2015 CMS report identified 384 PRTFs in the U.S., with nine operating in NC and 20 states with zero PRTFs.^12^ From 2008 to 2011, the federally-funded Community-Based Alternatives to PRTFs Demonstration project provided a Medicaid reimbursement waiver in nine states to provide services via a systems of care approach that enabled children eligible for PRTFs to receive care in their home and communities instead.^13^ The evaluation found the Demonstration “easily met cost-effectiveness tests and on average has consistently maintained or improved functional status for all children and youth.”^13,14^ Yet aside from this Demonstration evaluation, there are no known national studies of PRTFs. Even less is known about PRTFs’ effectiveness. The one recent systematic review of relevant peer-reviewed literature found mixed evidence of efficacy in improving child outcomes from generally low-quality studies.^15^

### Studies of PRTFs in NC

Two studies and one recent report examined PRTF use in NC. A 2012 study used Medicaid data to identify 2,157 youth admitted to PRTFs between 2008 and 2011.^16^ The number of youth served by PRTFs almost doubled in the study period. The study also sought to determine whether the profile of the youth population in PRTFs met the criteria established by the 2007 clinical coverage policy. Like NC’s current policy, the 2007 policy required children to demonstrate clinical need and that other less restrictive options were first tried and found unsuccessful. By analyzing Medicaid claims data, the authors concluded NC’s youth in PRTFs generally met these criteria: all had a psychiatric diagnosis, about half had a prior inpatient psychiatric hospitalization, and most had received other less restrictive forms of community-based treatment prior to entering a PRTF. However, the study did not explore factors associated with PRTF use or whether specific groups were more likely to enter PRTFs.

A 2015 study linked Medicaid claims data to other services databases to examine connections between youth’s use of PRTF, the income maintenance system, and child welfare services between 2007 and 2012.^17–19^ The study identified 2,730 youth from child welfare or income maintenance systems also admitted to PRTFs and, like the 2012 study, found that PRTF use rose during the study period. This study also yielded new findings about children’s experiences before, during, and after stays in PRTFs for the first time. First, recidivism was high: about one-third of children discharged later returned to a PRTF, on average 18 months after discharge. Second, medication use was very high among this group: about 85% of the youth in the sample were prescribed a psychotropic medication, 75% were prescribed an antipsychotic, and 66% were prescribed three or more psychotropic medications. Third, use of community-based services before and after care was surprisingly low given the youth’s high level of need. Only 50% had a prior claim for family therapy, 69% for individual therapy, and 42% had prior residential treatment. In multivariable models predicting PRTFs use, clinical need (particularly a trauma-related diagnosis) strongly predicted PRTF entry. Notably, contact with other public systems (specifically foster care) also emerged as a strong predictor of PRTF use. That is, while children entering PRTFs have high levels of mental health concerns, the association between system contact and PRTF entry suggests factors beyond clinical need may be driving PRTF utilization.

In 2020, the NC Department of Public Instruction, in conjunction with the NC Department of Health and Human Services’ Division of Mental Health, Developmental Disabilities, and Substance Abuse Services and Division of Health Service Regulation, provided a descriptive report to the NC General Assembly about the education of children in PRTFs.^20^ The report identified 15 contracts for educational services in 30 PRTFs. The contracted PRTFs reported educating 1,051 children, including 429 identified as “exceptional children” with an individualized education plan. The extent of the report’s assessment of child outcomes was its finding that 85% of children were discharged with an education plan in place (after an average 190-day length of stay). The report’s two recommendations were to improve collaboration between education and treatment staff and to improve interagency coordination.

### Aims of the Current Study

This study aimed to describe the population of children insured by NC Medicaid who received mental health services through PRTF placements. Two key indicators were assessed over time to examine recent utilization trends: (a) the proportion of children in PRTFs who were also in the public foster care system, and (b) the proportion of children placed in PRTFs out-of-state. We focus on these indicators because they represent aspects of service utilization which state agencies can potentially influence, as the state, through local social service agencies, assumes responsibility for the care of children in foster care. Thus, where children in foster care receive treatment for mental health concerns is the state and county government’s responsibility. Further, the licensing and availability of PRTFs within NC can be directly influenced by funding and regulatory decisions of the NC General Assembly and the NC Department of Health and Human Services. Beyond these two key indicators, we gathered descriptive information about the demographic, clinical, and service characteristics of children who entered PRTFs from 2015 to 2021. This data’s recency also enabled us to examine whether the COVID-19 pandemic was associated with any substantial changes in children placed in PRTFs.

## Methods

### Data and Study Population

This descriptive observational study relied solely on a limited data set of Medicaid enrollment and claims files provided by the NC Division of Health Benefits to the UNC Cecil G. Sheps Center for Health Services Research through the Carolina Cost and Quality Initiative. The secondary data files contain a census of Medicaid members and services in NC between calendar years 2015 and 2022. The sampling frame for this study was children and youth (ages five to 21 years) at the time of PRTF admission enrolled in NC Medicaid. Based on annual enrollment reports, the overall size of child NC Medicaid population was relatively stable during this time period.^21^ The study protocol was reviewed and determined to be exempt by the UNC Office of Human Research Ethics.

### Measures and Analysis

PRTF usage was identified using revenue center code 0911 or 0919 in institutional and professional claims. Since youth can be re-admitted to PRTFs, sometimes in the same calendar, and have varying lengths of stay, PRTF utilization was defined and calculated as an episode of care (EOC). A PRTF EOC was defined as 3 or more continuous days of care. PRTF stays separated by 30 or more days were counted as a new EOC. We calculated the annual number of unique children as well as a count of total EOCs. Before and during COVID-19 (defined as March 1, 2020) counts of children who received PRTF services were based on admission dates and children were described using demographic characteristics defined in Medicaid data: race (White, Black, American Indian, Asian/Pacific Islander, other); ethnicity (Hispanic, non-Hispanic); and gender (male, female). Diagnoses during the EOC were identified using *International Classification of Diseases, 9*^*th*^ *and 10*^*th*^ *Revisions*, ICD-9 (up to September 2015) and ICD-10 codes. Medications during the EOC for attention deficit hyperactivity disorder (ADHD) as well as anxiolytics, antidepressants, antipsychotics, and mood stabilizers were identified by *American Hospital Formulary Service* therapeutic classification code.

PRTF location (i.e., in- or out-of-state) was identified using billing provider state code to classify the provider location. Foster care service recipients were identified using enrollment data within 30 days of the beginning or end of the EOC indicating the specific Medicaid foster care program type. The aggregate net paid amount reimbursed for PRTF services by Medicaid each year was calculated. The net paid amount is much lower than the billed amount but represents the actual amount reimbursed. We adjusted the net paid amount for inflation using the Bureau of Labor Statistics Consumer Price Index 2021. Expenditure data is reported for a five-year period from 2018 to 2022 due to data quality concerns affecting reimbursement fields prior to 2018. All data management and analyses were performed in SAS Version 9.4 (SAS Institute, Cary, NC).

## Results

From 2015 to 2022, 10,966 PRTF episodes of cares were reimbursed for 10,038 children (duplicated) insured by NC Medicaid. Children averaged 14 years of age at admission every year. The average length of stay for a PRTF EOC ranged from 111 days (2015) to 131 days (2022). On average, 1,255 children were admitted to PRTFs over 1,370 EOCs each year. After peaking in 2018, the number of children and EOCs decreased each year through 2022. However, the proportion of children in foster care and placed out-of-state increased steadily. In 2022 the proportion of children in foster care increased steadily to 35% as did the proportion of PRTF placements out-of-state (44%). From 2018 to 2022, annual total net paid amounts ranged from $101 million to $125 million for a total five-year expenditure of $557 million (in 2021 dollars). From 2018 to 2022, the per-EOC expenditure increased from $67,000 to $104,000.

Table 1 compares the demographic characteristics, diagnoses, and select medications prescribed in PRTFs before and after March 2020. We observed slight increases in the proportions of PRTF admittees identified as female (43% to 45%), Black (36% to 38%), and American Indian (1.5% to 2.3%). Most diagnoses (DBD, psychotic disorder, substance use disorder) remained stable through pre- and peri-COVID periods. However, diagnoses for anxiety disorders (27% to 33% post-COVID), PTSD (25% to 30% post-COVID), and learning or intellectual disability disorders (1% to 3%) increased. From 2015 to 2022, we also observed an increase in the proportion of children who received ADHD medication and antidepressants. Notably, about 90% of PRTF EOCs included a prescription for an antipsychotic medication, although less than 3% had an associated psychotic disorder diagnosis.

**Table 1.** Demographic characteristics, diagnoses, and prescription drugs for episodes of care in PRTFs in North Carolina before and after March 2020

|  |  | Pre-COVID-19<br>(n = 7671)<br>Jan. 2015 to<br>Feb. 2020 |  | Peri-COVID<br>(n = 3295)<br>Mar. 2020 to<br>Dec. 2021 |  |
| --- | --- | --- | --- | --- | --- |
|  |  | n | % | n | % |
| Demographic Characteristics |  |  |  |  |  |
| Race | American Indian | 112 | 1.5% | 77 | 2.3% |
|  | Asian/Pacific Islander | 61 | 0.8% | 12 | 0.4% |
|  | Black or African American | 2768 | 36.1% | 1249 | 37.9% |
|  | White | 4589 | 59.8% | 1935 | 58.7% |
| Gender (Female) |  | 3284 | 42.8% | 1484 | 45.0% |
| Ethnicity (Hispanic) |  | 436 | 5.7% | 188 | 5.7% |
| Foster Care (Yes) |  | 2276 | 29.7% | 1259 | 38.2% |
| Diagnoses |  |  |  |  |  |
| ADHD |  | 1500 | 19.6% | 765 | 23.2% |
| Anxiety |  | 2053 | 26.8% | 1100 | 33.4% |
| PTSD |  | 1883 | 24.5% | 980 | 29.7% |
| Disruptive Behavior Disorder |  | 3459 | 45.1% | 1545 | 46.9% |
| Depression |  | 1216 | 15.9% | 606 | 18.4% |
| Psychotic Disorder |  | 205 | 2.7% | 76 | 2.3% |
| Substance Use Disorder |  | 242 | 3.2% | 107 | 3.2% |
| Learning or Intellectual Disability Disorder |  | 88 | 1.1% | 109 | 3.3% |
| Prescription Drugs |  |  |  |  |  |
| ADHD Medication |  | 3550 | 46.3% | 1707 | 51.8% |
| Antipsychotic Medication |  | 6761 | 88.1% | 2911 | 88.3% |
| Anxiolytics |  | 360 | 4.7% | 156 | 4.7% |
| Antidepressants |  | 4379 | 57.1% | 1963 | 59.6% |
| Mood Stabilizer |  | 3805 | 49.6% | 1441 | 43.7% |

## Discussion

NC, like the rest of the U.S., is experiencing a children’s mental health crisis. The American Academy of Pediatrics and other national groups recently declared a national emergency soon followed by an advisory from the U.S. Surgeon General. Facing a marked increase in service needs, states must decide *where* and *what type* of care children will receive. This analysis shows that prior to COVID and through the end of 2022, NC used PRTFs to provide care to over 1,000 children each year. Our analyses highlight three concerning trends that must inform NC’s ongoing response to the current crisis.

First, the increase in the proportion of children placed in out-of-state facilities (from 27% in 2016 to 44% in 2022) foregrounds potentially troubling systemic factors. This increase could reflect a decrease in in-state bed capacity or increases in lengths of stay, given the recently decreasing number of children placed in PRTFs annually. Alternatively, this trend could indicate that children placed in PRTFs increasingly have specialized care needs that can only be met by facilities with unique programming or specialties not available in NC. This might also reflect that NC only guarantees in-state treatment up to age 18, whereas Medicaid ensures care up to age 21.^22^ Regardless, out-of-state placements’ potential distance from children’s home communities, combined with PRTFs’ restrictiveness, could pose risks to children’s development of social connections and supports. Moreover, lack of direct oversight and legal jurisdiction in out-of-state PRTFs could create legal and logistical barriers to ensuring proper care. Further research should investigate the factors contributing to this increase in out-of-state placements and the potential for disparate outcomes between children placed within or outside of their home communities. Second, the overrepresentation of children in foster care within PRTFs is exceptionally concerning. In NC, about 9,000 children (ages 6-17 years) are in foster care annually,^23^ a relatively small proportion (< 1%) of children in the state.^24^ However, children in foster care constituted 26-42% of PRTF placements, depending on the year. This consistent, disproportionately high rate of PRTF placement for youth in foster care indicates that the capacity and effectiveness of community-based supports for system-involved youth are lacking. Although the prevalence of serious mental health challenges are higher among children in foster care, it is not clear that the overrepresentation can be explained by need alone.^8^ In fact, a study examining PRTF entries among children in NC with a maltreatment investigation from 2012 to 2017 found that children who entered foster care were over 10 times more likely to enter a PRTF, after controlling for other factors.^17^ The current study’s findings reinforce concerns that (a) residential treatment may be overutilized for the foster care population in order to provide structured placement options for children with placement instability and (b) children are possibly being removed to foster care to facilitate mental health treatment when their needs exceed the capabilities of their families and communities of origin. The many potential individual-, contextual-, and system-level drivers and decision-making processes potentially producing this glaring disparity are likely complex and demand further empirical examination and reform efforts. We also observed that anxiety and PTSD showed the largest point increases among youth in PRTFs comparing the prior and peri-COVID periods. This finding might be due to the increase in the proportion of youth in PRTFs also in foster care who have experienced child trauma related to abuse, neglect, and subsequent family separation. However, observed increases in the peri-COVID period in child anxiety and PTSD in the general population,^25^ and also among youth with pre-existing mental health conditions,^26^ is likely related to increases observed in PRTFs. Future research is needed to disentangle the effects of COVID-19 on existing stress and trauma-related disorders experienced by youth in foster care.

Third, our analysis shows an increase in psychotropic medication prescriptions and in internalizing disorders often associated with trauma (i.e., anxiety and PTSD). However, the fact that nine out of 10 PRTF placements included a prescription for antipsychotic drugs, the overwhelming majority did *not* include diagnosis for a psychotic disorder, corroborates prior suggestions that these medications are being overutilized potentially for general behavioral management. Some antipsychotic medications have been approved for use by youth diagnosed with certain conditions (Tourette’s syndrome, certain forms of bipolar disorder, and for irritability associated with autism spectrum disorder), at select ages.^27^ However, in many cases (e.g., for disruptive behavior disorder), antipsychotic medications are prescribed off-label.^28^ Child psychiatry experts have long warned that improperly prescribed antipsychotic medications might yield adverse side effects and negative outcomes.^29^

## Limitations

First, this study is descriptive, and we do not suggest PRTF stays are caused by any youth characteristics, or vice versa. Second, “out-of-state” as an indicator of a youth’s distance from their home community is somewhat crude. For example, some youth whose communities are near the borders of other states may be closer to home at an out-of-state facility than at another facility located in a distant part of NC. Third, diagnosis codes in claims data are entered for reimbursement purposes and diagnosis codes appearing in these claims might not reflect the underlying disorder. Similarly, claims for prescribed drugs do not reflect that the drugs were taken, and it is not possible to know why any medication was prescribed.

## Conclusion

Ideally, youth with behavioral health needs will be treated in their communities, receiving individual and family services from nearby outpatient clinics, and in many cases supplemented by appropriate medications with proven safety and efficacy. As a last resort, some youth with more substantial mental and behavioral health needs may need a more intensive level of care, potentially supervised in a PRTF. PRTFs have the potential to offer appropriate, high-quality care, but serious concerns remain about the effectiveness of care delivered in PRTF settings and about the safety and well-being of children in low-quality PRTFs.^11,15^ PRTFs will likely continue to be a critical provider in the children’s mental health service system, particularly given the alarming trends in children’s mental health needs in the post-COVID era. Yet given these trends, policymakers and stakeholders should ask more questions about the quality of care provided in PRTFs and demand accountability where care is inadequate.

**Figure 1.**
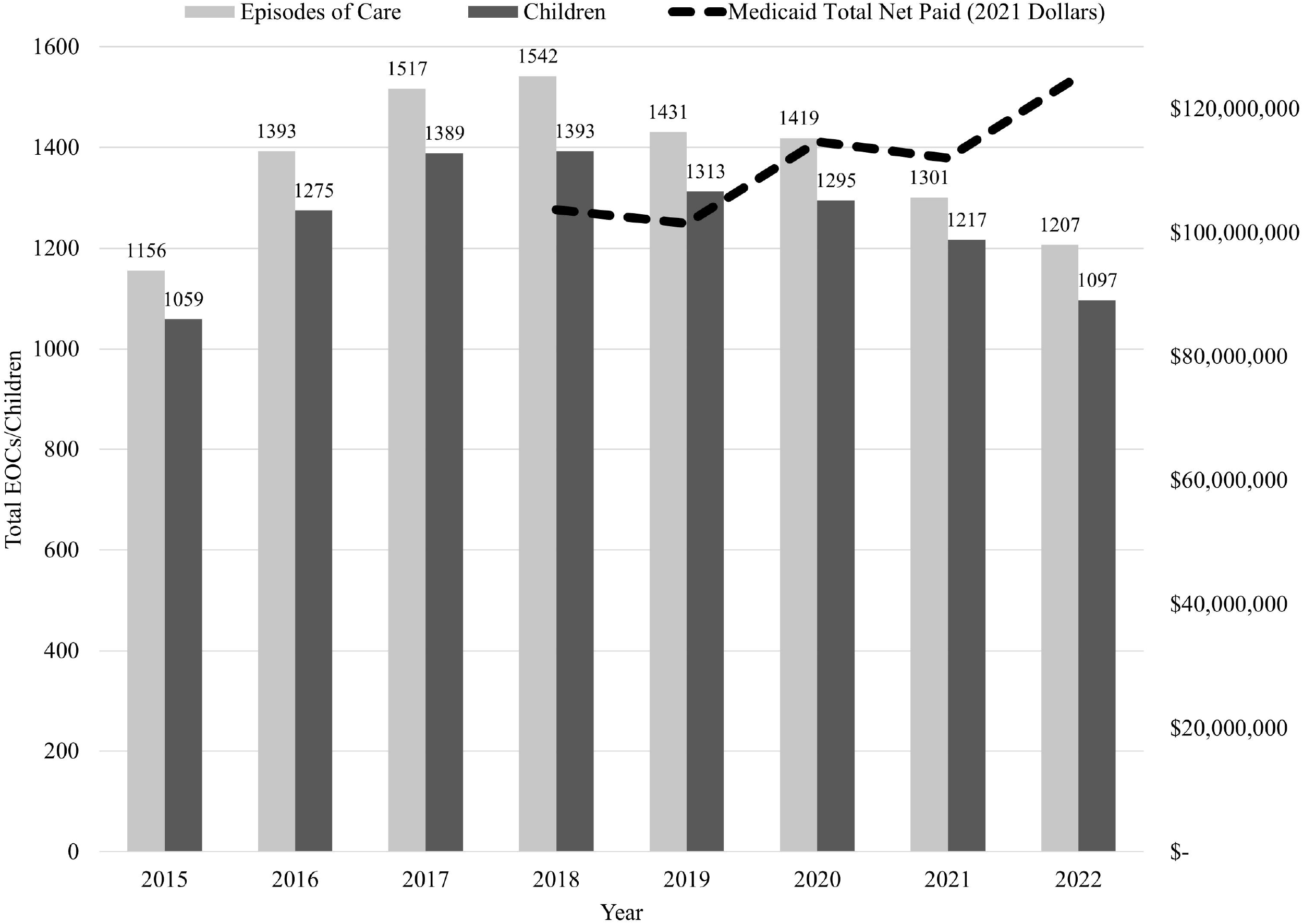
Annual Counts of Children, Episodes of Care, and Expenditures for PRTFs in North Carolina.

**Figure 2.**
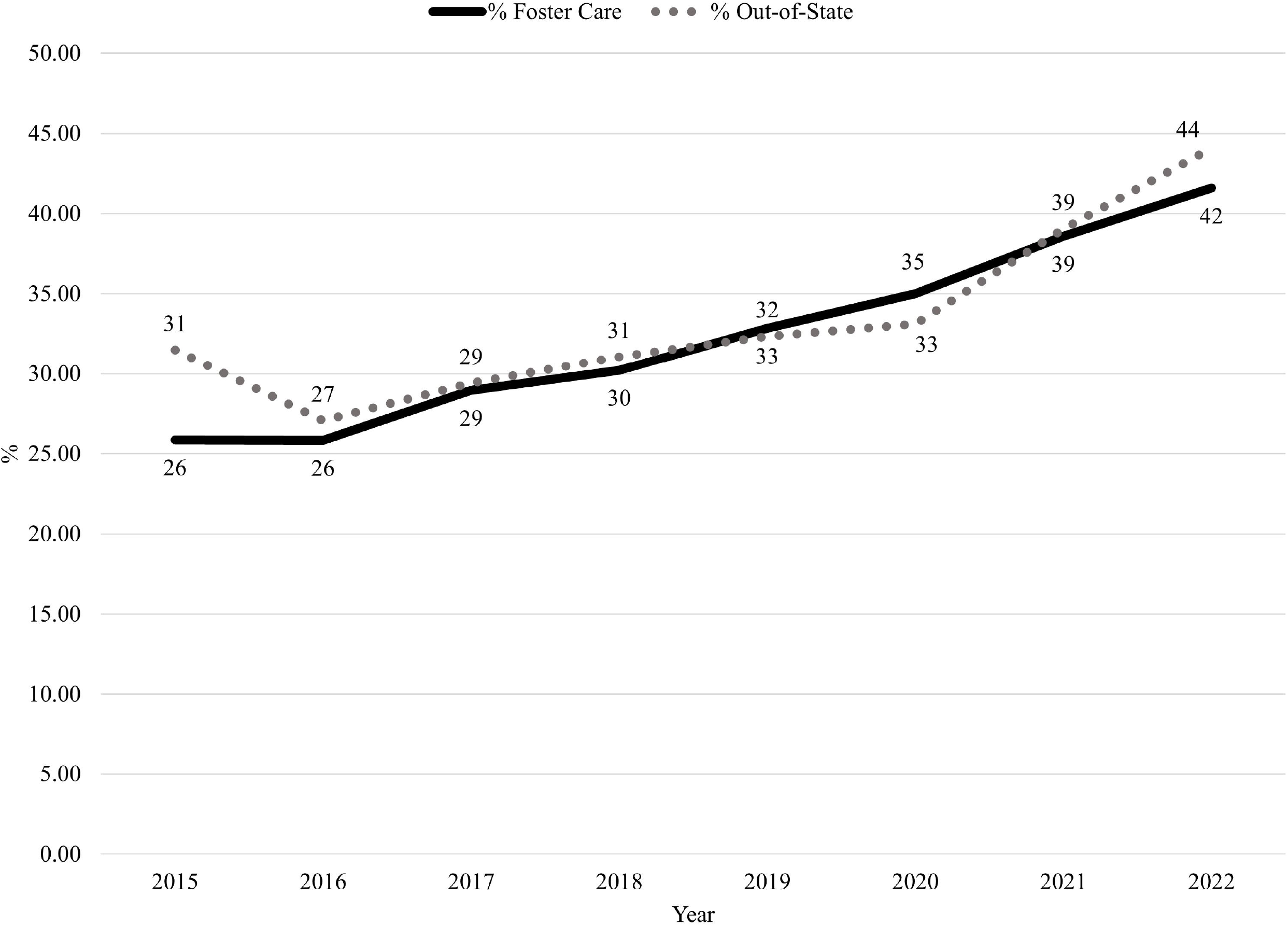
Proportion of NC Children in PRTFs in Foster Care and Out-of-State Increasing over Time

## Data Availability

Data used for this study is not publicly available.

## Acknowledgements

We would like to acknowledge partners at the North Carolina Division of Child and Family Well-Being and the North Carolina Division of Health Benefits who provided input and expertise on this study. The database infrastructure used for this project was funded by the Cecil G. Sheps Center for Health Services Research; the Department of Health Policy and Management; UNC Gillings School of Global Public Health; the CER Strategic Initiative of UNC’s Clinical and Translational Science Award (UL1TR001111); and the UNC School of Medicine.

